# Patient Perspectives on Buprenorphine Treatment for Opioid Use Disorder and Preferences for Long-Acting Injectable Formulations: Findings from a National Online Survey

**DOI:** 10.64898/2026.02.05.26345663

**Authors:** Tyler S. Oesterle, Nicholas L. Bormann

**Affiliations:** Department of Psychiatry and Psychology, Mayo Clinic, Rochester, MN, USA

**Keywords:** opioid use disorder, buprenorphine, long-acting injectable, depot buprenorphine, patient preferences, patient-reported outcomes, shared decision-making

## Abstract

**Background:** Long-acting injectable buprenorphine (LAIB) has been positioned as a potentially transformative option for opioid use disorder (OUD), in part because patient experiences reported in qualitative studies emphasize reduced daily burden, increased “freedom,” reduced stigma, and fewer pressures related to diversion—while also noting barriers such as insufficient information, early adverse experiences, and concerns about coercion.

**Methods:** We conducted a cross-sectional online survey of adults recruited from the Behavioral Health Research Panel (BHRP). Eligibility included age ≥18, English literacy, and OUD diagnosis or problematic opioid use within the past 5 years. Survey content assessed buprenorphine experience, knowledge and attitudes toward LAIB, attribute preferences, and open-text feedback. Descriptive statistics were generated; analyses were stratified by buprenorphine experience (experienced vs naïve).

**Results:** Among 105 participants, 82.9% reported prior buprenorphine use, and 17.1% were buprenorphine-naïve. Overall, 53.3% preferred a long-acting injection regimen (weekly/monthly/3-monthly) versus 46.7% preferring a daily oral tablet/film. Convenience and adherence-related themes (e.g., not missing doses, fewer visits) drove LAIB preference, while oral-route preference and concerns about side effects and safety were prominent among those favoring oral formulations.

**Conclusions:** In this national convenience sample, preferences were nearly evenly split between daily oral and long-acting injectable buprenorphine regimens, with a slight overall preference for LAIB. Findings align with the qualitative literature, emphasizing the practical and psychosocial benefits of LAIB, alongside persistent needs for improved education, shared decision-making, and attention to tolerability, safety perceptions, and cost/coverage barriers.

## Introduction

Opioid use disorder (OUD) remains a major public health challenge, and buprenorphine is an evidence-based medication treatment delivered most commonly via daily sublingual tablets or films. Long-acting injectable buprenorphine (LAIB; “depot” or “extended-release” formulations) was developed to reduce the day-to-day demands of dosing and to address adherence challenges inherent to chronic conditions. Patient-centered perspectives are critical when evaluating these formulations because acceptability, perceived fit, and treatment burden strongly influence uptake and continuation. (10, 4)

Qualitative interviews with patients receiving depot buprenorphine highlight **social and practical benefits**, including increased freedom to participate in work and family life and psychological benefits such as “feeling normal.” These studies also describe **early challenges**, including withdrawal-like symptoms early in treatment, and emphasize that **trust and reliable information** from clinicians can help patients navigate initial difficulties and support persistence. In another qualitative study focused on patients with ongoing substance use and psychiatric comorbidities, participants described **social benefits, shifts in identity and self-perception**, and relief from perceived pressures to divert medication, while also reporting that **pre-treatment information was sometimes insufficient** and that experiences of **coercion** were linked to negative attitudes toward depot medication. (5,6)

Longitudinal qualitative work further suggests LAIB may influence patients’ interpersonal contexts: during the first year of LAIB treatment, relationship supports (family, children) can motivate treatment, while social isolation and variable contact with services may shape patient experience over time. Complementing qualitative findings, patient-reported outcomes from an open-label randomized trial demonstrated **higher treatment satisfaction** among individuals receiving weekly or monthly depot buprenorphine compared with daily sublingual buprenorphine, supporting satisfaction as a meaningful endpoint when comparing OUD medications. (4,7)

### Why this survey matters

While qualitative studies provide depth into lived experience, broader surveys can quantify preferences and identify population-level barriers and facilitators. We therefore conducted a national online survey to assess patient perspectives on buprenorphine therapies, focusing on (1) lived experience with buprenorphine (oral and LAIB), (2) regimen preferences, (3) perceived benefits and concerns regarding LAIB, and (4) desired improvements in buprenorphine treatments.

## Methods

### Study design and setting

This was a cross-sectional online survey conducted among adults recruited from the Behavioral Health Research Panel (BHRP).

### Data source: Behavioral Health Research Panel (BHRP)

BHRP is a voluntary research network of individuals who opted in to be contacted for research after completing the Addiction Severity Index - Multimedia Version (ASI - MV) assessment (1-3).

### Eligibility criteria

Participants were eligible if they: (1) were ≥18 years old, (2) could read and understand English, (3) were BHRP panelists, and (4) reported either an OUD diagnosis or problematic opioid use (heroin, street fentanyl, or prescription opioids) within the past 5 years.

### Recruitment and procedures

Panelists received an email invitation containing a link to an online screener and electronic informed consent. The survey was administered using SurveyMonkey. Participants were offered a $20 electronic gift card upon completion (email address optional).

### Survey measures

Survey domains included: demographics; substance use history and recency; buprenorphine use history (oral and LAIB); settings of initiation; satisfaction; willingness to switch from oral to LAIB; perceived benefits/concerns; regimen preferences; and open-text improvement suggestions.

### Definitions

Participants were classified as **buprenorphine experienced** if they reported any prior oral or LAIB use for OUD, and **buprenorphine naïve** if they reported no prior buprenorphine use.

### Human subjects protections

The study was reviewed and approved by WCG Institutional Review Board (IRB protocol 20250574; approved 24 February 2025).

### Statistical analysis

Descriptive statistics were generated, including stratification by buprenorphine experience group. Results for small subgroups (<30) were interpreted cautiously, consistent with the study report.

## Results

### Participant disposition and sample size

Survey data were collected from **25 February 2025 through 24 March 2025**. The screener was accessed 175 times; 105 unique completed surveys were included after excluding incomplete/duplicate responses (n=5).

### Participant characteristics

Among 105 participants, **63.8% were female**, the mean age was **39.2** (range 23–74), and **87.6% identified as White**. Most resided in the US South (71.4%), with respondents reporting urban (35.2%), suburban (29.5%), and rural (27.6%) settings.

### OUD diagnosis and opioid use history

Most participants reported a lifetime OUD diagnosis (83.8%). Problematic use in the prior 5 years was commonly reported for prescription opioids (74.3%), heroin (59.1%), and street fentanyl (54.3%).

### Buprenorphine experience and formulations used

Overall, **82.9%** were buprenorphine-experienced, and 17.1% were buprenorphine-naïve. Oral buprenorphine experience was common; smaller proportions reported LAIB experience. Among buprenorphine-experienced participants, **16.1%** reported Sublocade® monthly use and **5.8%** reported Brixadi® monthly use (with 1.2% reporting Brixadi® weekly).

### Willingness to switch from oral to LAIB

Among participants currently taking oral buprenorphine, **56.9%** indicated they would consider switching to LAIB. Among those not willing to switch, common reasons included preferring oral medication, concerns about side effects, and concerns about the injection dose amount.

### Satisfaction with buprenorphine

Most oral buprenorphine users reported at least moderate satisfaction. Among LAIB users, the majority of Sublocade® monthly and Brixadi® monthly users reported at least moderate satisfaction, while the single weekly Brixadi® user reported low satisfaction.

### Preferences for ideal regimen

Overall preference was **53.3%** for a long-acting injection regimen versus 46.7% for a daily oral tablet/film. The most selected option was a **3-month injection (27.6%)**, followed by daily film (24.8%), daily tablet (21.9%), monthly injection (20.0%), and weekly injection (5.7%). Preferences differed by experience group: buprenorphine-experienced participants more often preferred LAIB (55.2%), whereas buprenorphine-naïve participants more often preferred oral regimens (55.6%).

### Reasons for preferring LAIB vs oral therapy

Reasons for preferring LAIB included not worrying about missing a daily dose, fewer office visits, longer duration of action, less risk of misplacing medication, and greater stability. Reasons for preferring oral formulations included preference for the oral route, worries about side effects, injection safety, and fear of needles.

### Open-text themes: “How could buprenorphine be better?”

Common improvement themes included: **side effects** (including dental concerns for oral formulations), **ease of induction/tapering, taste, cost, availability**, and concerns about withdrawal or precipitated withdrawal—particularly in the context of fentanyl exposure

## Discussion

In this national online convenience sample, respondents expressed **substantial interest in long-acting injectable buprenorphine**, with a modest overall preference for LAIB regimens compared with daily oral formulations. Convenience and adherence-related benefits (fewer missed doses, fewer visits) were among the most commonly endorsed reasons to preferring LAIB, mirroring recurring themes in qualitative patient interviews, in which depot formulations are associated with improved day-to-day functioning and “freedom” from daily dosing rituals (5,6).

Notably, patient experience studies emphasize that the transition to LAIB is not uniformly positive: early adverse symptoms, perceived inadequate effect, and concerns about insufficient information can shape discontinuation or refusal. This aligns with our findings that among LAIB users, common dislikes included injection pain, feeling the medication did not work, side effects, and safety concerns; among oral users hesitant to switch, worries about side effects and injection-related concerns were frequent (6,6).

Another consistent point of convergence with PubMed-indexed qualitative literature is the centrality of **information quality and shared decision-making**. Patients interviewed in depot buprenorphine qualitative studies described the importance of trustworthy clinician relationships and adequate pre-treatment counseling, while coercion or a negative “pharmaceutical atmosphere” undermined acceptability. In our survey, open-text responses similarly stressed the need for better counseling about side effects, tapering, and induction—particularly around withdrawal and precipitated withdrawal, a salient concern in fentanyl contexts (5,6)

Our results also complement evidence from patient-reported outcomes trials. In an open-label randomized trial, patients receiving depot buprenorphine reported higher satisfaction than those receiving daily sublingual buprenorphine, supporting satisfaction and reduced treatment burden as meaningful endpoints. Consistent with this, survey participants cited expected benefits such as a stable effect and reduced need for frequent visits, and LAIB users endorsed practical advantages such as avoiding medication misplacement and fewer office visits (4)

Finally, the finding that **over half of buprenorphine-naïve participants were unaware that buprenorphine was available** underscores the importance of education and access pathways. This is especially relevant given qualitative work showing that perceived access constraints may influence decision-making and lead to a sense of needing to “rush” decisions when depot options are scarce (7)

## Limitations

This survey used a **convenience sample** of BHRP panelists and may not generalize to all people with OUD. All measures were self-reported and not verified by biospecimens or medical records. Some subgroup analyses were based on small cell sizes (e.g., LAIB users), limiting the ability to draw inferences.

## Conclusions

In this national survey of adults with OUD diagnosis or problematic opioid use history, preferences were nearly evenly split between daily oral and long-acting injectable buprenorphine regimens, with a slight overall preference for LAIB. Participants valued LAIB for adherence support and reduced treatment burden, while concerns centered on injection pain, safety perceptions, side effects, and cost/coverage barriers. Together with PubMed-indexed qualitative evidence, these findings support implementation strategies emphasizing shared decision-making, high-quality patient education, and attention to tolerability and access barriers.

## Data Availability

All data produced in the present study are available upon reasonable request to the authors

